# Characterizing Left Atrial Failure via the Atrial Booster Preload-Performance Relationship

**DOI:** 10.64898/2026.02.13.26346251

**Authors:** Doron Aronson, Ida Maiorov, Sobhi Abadi, Jonathan Lessick

**Author notes:** **Address for Correspondence:** Doron Aronson, MD Department of Cardiology, Rambam Medical Center, POB 9602, Haifa 31096, Israel.

## Abstract

**Background:** Left atrial (LA) remodeling, a hallmark of chronically elevated LA pressure, is characterized by enlargement and functional impairment. While global and reservoir LA functions are well described, the role of LA booster function and its failure remains poorly defined.

**Objectives:** To characterize LA booster function using cardiac computed tomography angiography (CCTA) and to evaluate the relationship between LA preload, booster performance, remodeling, and clinical outcomes.

**Methods:** We retrospectively analyzed 975 patients who underwent spiral CCTA between 2010 and 2018. Phasic LA and LV volumes were obtained, from which LA reservoir and booster functions were derived. LA performance curve was constructed by plotting LA pre-A volume (preload) against LA booster stroke volume. Clinical outcomes (heart failure, stroke, or cardiovascular death) were analyzed based on the LA performance curve.

**Results:** LA pre-A volume strongly correlated with LA end-systolic volume (r=0.92, p<0.001). The LA booster stroke volume displayed an inverted U-shaped relation to LA pre-A volume (linear coefficient 0.64, P<0.0001; squared coefficient-0.0029, P<0.0001). The atrial booster function curve reached its vertex at 107 mL (95% CI 90 to 113 mL), indicating that the booster pump response for the increased preload is exhausted at this point. Booster dysfunction was associated with impaired reservoir function (r=0.77, p<0.001) and reduced LA systolic flow rates (-0.79, *P*<0.001). Patients with increased LA pre-A volume but reduced booster volume (“LA failure”) exhibited the highest event rate of the combined endpoint of heart failure, stroke or cardiovascular mortality (43.2%, 95% CI 33.6–54.2%).

**Conclusions:** LA enlargement predominantly serves to increase LA pre-A volume to sustain booster function. LA contractile dysfunction affects global LA function via a concomitant reduction in LA reservoir volume. LA failure can be defined as reduced booster contraction despite elevated preload, portending poor clinical outcomes.

## Background

The left atrium (LA) plays a central role in maintaining cardiovascular homeostasis and assumes greater importance in the setting of heart failure (HF) (1). Elevation in LA pressure leads to structural and functional remodeling, characterized by LA dilatation and impaired atrial function (2,3). The extent of LA remodeling correlates with the severity of diastolic dysfunction and contributes to symptoms (4,5) and disease progression (6), and is generally regarded as a maladaptive process.

In clinical practice, LA failure is typically defined by measures of global LA function—such as total emptying fraction (6–11)—or reservoir function, including LA reservoir strain and LA expansion index (9,12,13). More broadly, LA failure encompasses any form of atrial impairment—anatomical, mechanical, or electrical—that leads to reduced cardiac performance and symptoms (14).

During atrial systole, LA contraction augments LV end-diastolic fiber length and tension, enhancing subsequent ventricular contraction. Accordingly, a reduction in LA booster function results in increased mean LA pressure and attenuates LV contractility (15). Despite its physiological importance, data on the role of LA booster function in LA failure remain limited.

Cardiac computed tomography angiography (CCTA) allows automated, voxel-based quantification of phasic LA volumes (16,17), offering high spatial resolution and improved sensitivity for detecting subtle alterations in LA remodeling and function. In the present study, we used CCTA to evaluate LA booster function and its relationship to LA remodeling and other phasic indices, particularly LA pre-A volume—a measure of LA preload. We demonstrate that the LA performance curve, relating booster preload to stroke volume, can be used to characterize the spectrum of LA dysfunction.

## Methods

The requirement for written informed consent was waived by the Rambam Hospital Institutional Review Board that gave its approval for this study. From our database of patients studied for the evaluation of LA function by CT (11), we identified patients who had undergone both a cardiac CT examination using spiral scanning with retrospective gating. We excluded patients with chronic atrial fibrillation, unstable patients and insufficient CT quality. Retrospective data were analyzed using fully automatic segmentation of the heart chambers, producing phasic volume curves of each heart chamber. LA volume–based indices were calculated as previously described (18,19).

For each patient, the time-volumes curves were used to identify LV end-diastolic volume (LVEDV), LV end-systolic volume (LVESV), LA maximum end-systolic volume (LAESV), LA pre-A volume and LA minimum end-diastolic volume (LAEDV) as well as LV mass (LVM) at end-diastole. LV ejection fraction (LVEF) and measures of atrial function were subsequently calculated.

Global, passive and active LA function were evaluated by total emptying fraction (LAEF**_Total_**), passive LAEF (LAEF**_Passive_**) and active LAEF (LAEF**_Booster_**), respectively. These fractional volume changes were calculated as follows (20,21):

- LAEF_Total_ = 100*(LAVmax-LAVmin)/ LAVmax
- LAEF_Passive_ = 100* (LAVmax-LAVpre-A)/LAVmax
- LAEF_Booster_=(LAVpre-A-LAVmin)/ LAVpre-A.

Measures of left atrial reservoir function included LA reservoir volume (LAV**_max_** - LAV**_min_**), LAEF**_Total_** and LA expansion index [LAEI = 100*(LAV**_max_** - LAV**_min_**)/LAV**_min_**)], representing the relative LA volume changes during the reservoir phase of LA function (9,19). An LA performance curve was constructed, relating preload measured as LAV**_pre-A_** to LA booster performance, measured as LA booster volume change (akin to a Frank-Starling curve).

Atrial emptying was divided into 3 components: 1) early “passive” emptying volume; 2) a late “active” (booster) phase; and 3) Conduit volume, defined as the volume of blood that passes through the LA that cannot be accounted for by early or booster pump function: LV stroke volume - LA reservoir volume (6). These parameters were expressed in terms of their percent contribution to left ventricular stroke volume.

In a subset of 79 patients randomly selected from the cohort, LA phasic volume change was calculated for each cardiac phase (n=20) throughout the cardiac cycle. The mean duration of each phase was 47 ± 8 ms, depending on the patient’s heart rate. From the LA volume-time curve and its first derivative, we calculated the peak slope (representing flow rate) of LA filling during the reservoir phase and of LA emptying during the atrial booster phase.

### Statistical analysis

Continuous variables are presented as mean±SD or medians (with interquartile ranges), and categorical variables as numbers and percentages. Baseline characteristics of the groups were compared using analysis of variance for continuous variables and by the χ**^2^** statistic for categorical variables. Basic geometric parameters measured by echo and by CT, such as LA volumes were compared by Pearson correlation. Relations of various parameters with the atrial flow rate were expressed in terms of linear regression correlation coefficients.

The LA Frank-Starling curve was constructed by plotting the LA pre-A wave volume and atrial kick volume. The relationship was assessed as a continuous function and analyzed with a quadratic regression model. The quadratic term was chosen to allow for some curvature in the relationship. The quadratic equation provided a significantly better fit than did linear regression (the coefficient of the quadratic equation was significantly different from 0). The vertex (turning point) of the quadratic curve was calculated as-β1/β2, where β1 is the coefficient of the linear term and β2 is the coefficient of the quadratic term.

The relationship between pre-A volume and atrial kick volume was also explored using a nonparametric regression method with a Gaussian Kernel function to estimate potentially nonlinear response function, with CIs obtained using 1000 bootstrap samples. Quantile regression was also used to explore the same relationship.

Differences were considered statistically significant at the 2-sided P<0.05 level. Statistical analyses were performed using STATA Version 18.0 (College Station, TX).

## Results

The study cohort comprised of 1067 patients in sinus rhythm, undergoing spiral cardiac CT from January 2010 to June 2018. We excluded 42 patients due to non-sinus rhythm or poor CT quality. Fifty additional patients were excluded due to missing full LA function data. Full volumetric analysis of LA function was performed in the remaining 975 patients (mean age 64 ± 14, 390 females).

### LAV_pre-A_ and atrial booster volume curve

Figure 1 depicts the relationship between LAV**_pre-A_** and atrial booster volume. The relationship followed an inverted U relationship, such that the volume of the atrial kick initially increased with increasing preload (LAV**_pre-A_**) but progressively decreased at higher LAV**_pre-A_** (Figure 1; linear coefficient 0.64, *P*<0.0001; squared coefficient-0.0029, *P*<0.0001). The active atrial function curve reached its vertex at 107 mL (95% CI 90 to 113 mL), indicating that the booster pump response for the increased preload is exhausted at this point in some individuals, and declines despite higher booster pump preloads. Nonparametric modeling (Supplemental figure 1) and quantile regression (Supplemental Figure 2) of the relationship between LA pre-A volume and LA kick volume showed a similar inverted U-shaped pattern.

**Figure 1:**
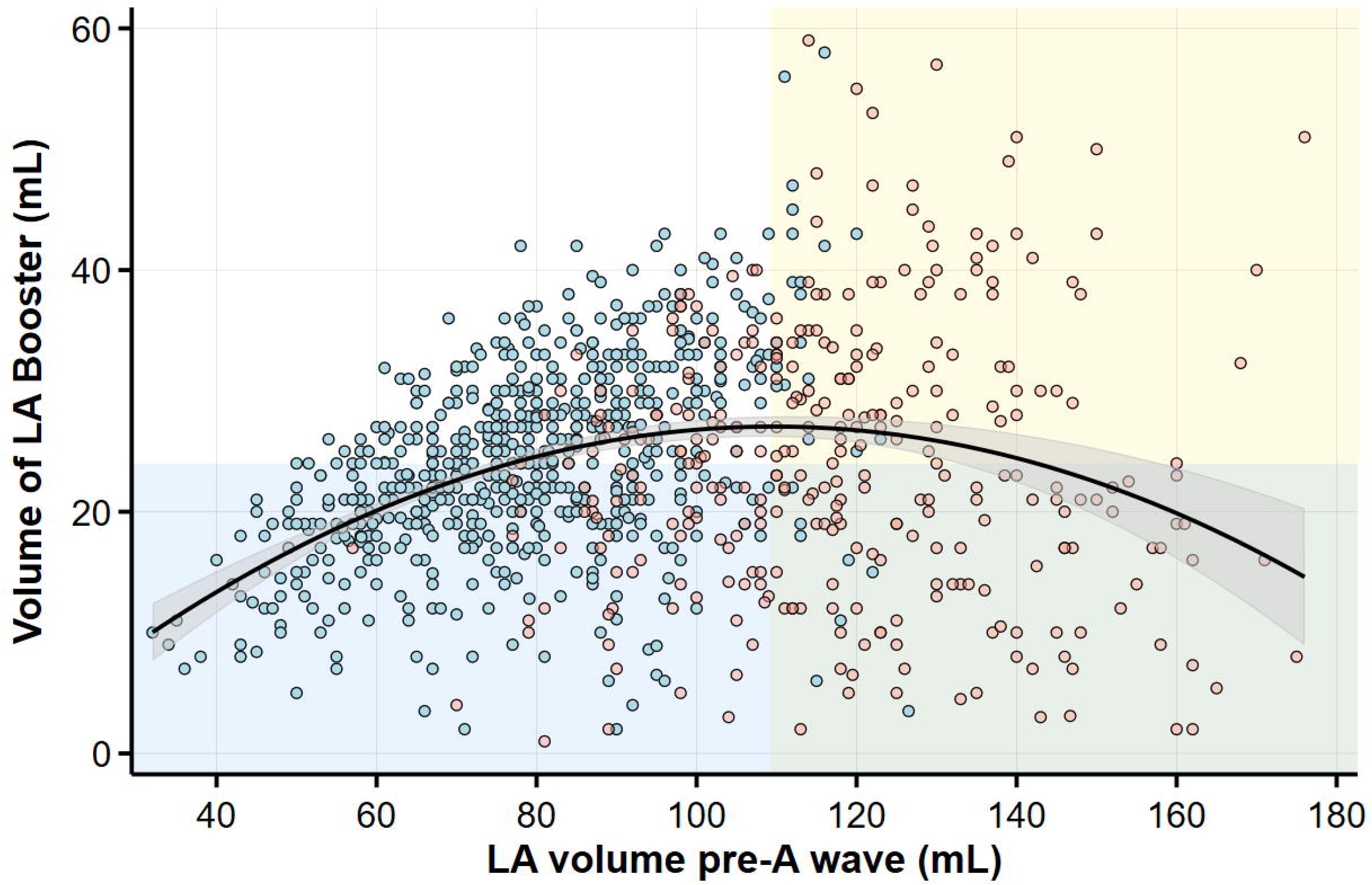
Relationship between the pre-A wave LA volume and the atrial kick volume. The fitted line is a quadratic function. The blue and orange dots denote normal and increased left atrial volume, respectively.

However, there was a substantial variability in the LA response to increased atrial preload. In some patients the volume of the atrial kick continues to increase with high LAV**_pre-A_** values and displays adequate and robust atrial response to increased preload. Other patients demonstrated a reduction in the atrial kick volume even at relatively low LAV**_pre-A_**, indicating that in some patients, the atrial booster function continues to increase at high atrial pre-A volumes, whereas in others it rapidly declines. Of note, an increase in LA booster volume occurred almost exclusively in patients with LA enlargement (Figure 1).

A similar inverted U-shape pattern was observed in patients with preserved ejection fraction and patients with mildly reduced or reduced LV ejection fraction (supplemental Figure 3) and in men and women (supplemental Figure 4).

Based on the LA pre-A volume and the LA booster volume, 4 different groups were created: Group 1 (LAVpre-A < 110 mL and LA booster volume < 25 mL), Group 2 (LAVpre-A < 110 mL and LA booster volume ≥ 25 mL) the reference group, Group 3 of patients with appropriate booster response to the increased booster preload (LAVpre-A ≥ 110 mL and LA booster volume ≥ 25 mL), and Group 4 of patients with reduced booster function despite increased booster preload (LAVpre-A ≥ 110 mL and LA booster volume < 25 mL), indicating LA failure.

The clinical characteristics of the 4 groups are shown in Table 1. Compared to patients in group 3, Patients in group 4 were older, more likely to be women, hypertensive, and with a history of myocardial infarction. The CTCA findings of the 4 groups are shown in Table 2. Patients in group 4 had lower LVEF, larger LV volume and higher LVMI. Patients in group 3 had reduced measures of early passive emptying and preserved measures of LA booster function, while patients in group 4 had a reduction in both early passive and booster functions. Although the booster function in group 3 was preserved, this was achieved via a marked increase in LA pre-A volume. However, even patients in group 3 had some reduction in global LA function (LAEF**_Total_**).

**Table 1:** Baseline Clinical Characteristics.

| **LA pre-A Volume** | **<110 mL** |  | **≥110 mL** |  |  |
| --- | --- | --- | --- | --- | --- |
| **Atrial kick volume** | **<25 mL** | **≥ 25 mL** | **≥25 mL** | **< 25 mL** | ***P* value** |
| **Group** | **1** | **2** | **3** | **4** |  |
| Age (years) | 61 ± 15 | 64 ± 13 | 68 ± 13 | 71 ± 13 | <0.0001 |
| Male gender | 204 (51) | 105 (31) | 55 (43) | 31 (23) | <0.0001 |
| Body surface area (m<sup>2</sup>) | 1.8 ± 0.2 | 2.0 ± 0.2 | 2.0 ± 0.2 | 1.9 ± 0.2 | <0.0001 |
| Hypertension | 246 (61) | 237 (71) | 106 (80) | 96 (75) | <0.0001 |
| Diabetes Mellitus | 145 (36) | 111 (33) | 46 (35) | 52 (41) | 0.52 |
| Coronary artery disease | 143 (35) | 144 (43) | 64 (48) | 60 (47) | 0.02 |
| Previous CABG | 84 (21) | 85 (25) | 36 (27) | 33 (26) | 0.31 |
| Previous MI | 89 (22) | 110 (33) | 36 (27) | 35 (27) | 0.01 |

Values are presented as n (%), mean ± SD or median (interquartile range).

**Table 2:** Baseline CT Characteristics.

| **LA pre-A Volume** | **<110 mL** |  | **≥110 mL** |  |  |
| --- | --- | --- | --- | --- | --- |
| **Atrial kick volume** | **<25 mL** | **≥ 25 mL** | **≥25 mL** | **< 25 mL** | ***P value*** |
| LVEDVI (ml/m<sup>2</sup>) | 76 [66–89] | 79 [72–91] | 97 [79–107] | 103 [83–134] | <0.0001 |
| LVESVI (ml/m<sup>2</sup>) | 30 [21–49] | 27 [21–37] | 37 [24–49] | 47 [30–78] | <0.0001 |
| Stroke volume (mL) | 85 ± 25 | 101 ± 19 | 114 ± 26 | 98 ± 27 | <0.0001 |
| LVEF (%) | 60 [51–69] | 66 [58–72] | 61 [52–69] | 54 [39–63] | <0.0001 |
| LVMi (g/m<sup>2</sup>) | 67 [56–79] | 73 [64–87] | 98 [79–122] | 94 [75–115] | <0.0001 |
| LAESVI (ml/m<sup>2</sup>) | 50 [42–59] | 54 [48–60] | 70 [65–81] | 79 [70–89] | <0.0001 |
| **Global LA Function** |  |  |  |  |  |
| LATEF (%) | 40 [32–46] | 47 [42–51]* | 37 [32–42] | 19 [14–25] | <0.0001 |
| **Reservoir function** |  |  |  |  |  |
| Reservoir volume index (ml/m<sup>2</sup>) | 19 [16–23] | 25 [22–28] * | 26 [22–28] | 16 [12–20] | 0.0001 |
| LA expansion index (%) | 67 [48–85] | 88 [73–103]* | 60 [48–63] | 26 [19–35] | <0.0001 |
| **Early passive function** |  |  |  |  |  |
| LA passive emptying fraction (%) | 18 [13–23] | 17 [13–20] | 12 [19–15] | 10 [7–13] | <0.0001 |
| LA passive volume (mL) | 16 [11–22] | 17 [12–22] | 16 [13–22] | 15 [10–20] | 0.02 |
| Early passive contribution to LV filling (%) | 19 [14–26] | 17 [13–23] | 15 [11–19] | 16 [11–20] | 0.0001 |
| **Booster function** |  |  |  |  |  |
| LA Pre-A Volume index (ml/m<sup>2</sup>) | 40 [33–49] | 45 [40–50] | 62 [57–71] | 71 [63–80] | <0.0001 |
| LA active emptying fraction (%) | 21 [17–25] | 29 [26–33] | 24 [21–28] | 11 [7–14] | <0.0001 |
| LA Booster Volume | 19 [15–22] | 30 [27–33] | 34 [29–40] | 16 [10–20] | <0.0001 |
| Booster contribution to LV filling (%) | 22 ± 8 | 31 ± 6* | 33 ± 11 | 16 ± 22 | <0.0001 |

Values are presented as median (interquartile range)

**Table 3:** Cox proportional hazards models for the admission for the composite endpoint of heart failure, stroke and cardiovascular mortality.

| LA functional parameters | Univariable |  | Multivariable* |  |
| --- | --- | --- | --- | --- |
|  | HR (95% CI) | *P* value | HR (95% CI) | *P* value |
| **Pre-A Vol<110 / Atrial Kick Vol<25** | 1.37 (0.90–2.13) | 0.16 | 1.47 (0.94–2.30) | 0.09 |
| **Pre-A Vol<110 / Atrial Kick Vol≥25** | 1.0 (Referent) |  | 1.0 (Referent) |  |
| **Pre-A Vol≥110 / Atrial Kick Vol≥25** | 3.21 (1.97–5.25) | <0.0001 | 1.95 (1.18–3.25) | 0.01 |
| **Pre-A Vol≥110 / Atrial Kick Vol<25** | 4.07 (2.52–6.57) | <0.0001 | 2.43 (1.47–4.00) | 0.001 |

\* Adjusted for age, sex, hypertension, diabetes mellitus, coronary artery diseases, coronary artery bypass surgery, left ventricular ejection fraction, valvular heart disease, and left atrial volume index.

#### LA booster flow rates

The flow rates into the LV during LA systole were calculated in a subgroup of 74 patients. Based on these data we constructed time-volume curves of the 4 study groups (Figure 2). Compared to Group 2 patients with high booster volume and low pre-A volume (booster flow rate 216 mL/s, 95% CI 197 to 236 mL/s), similar booster flow rate was seen similar in Group 3 patients with high booster and high pre-A volumes (196 mL/s, 95% CI 172 to 219, *P*=0.19), but lower flow rates were obtained in Group 1 patients with low booster volumes and low pre-A volumes (98 mL/s, 95% CI 80 to 116, *P*<0.001) and markedly lower rates in group 4 patients with low booster volume and high pre-A volume (81 mL/s; 95% CI 57 to 105, *P*<0.001).

**Figure 2:**
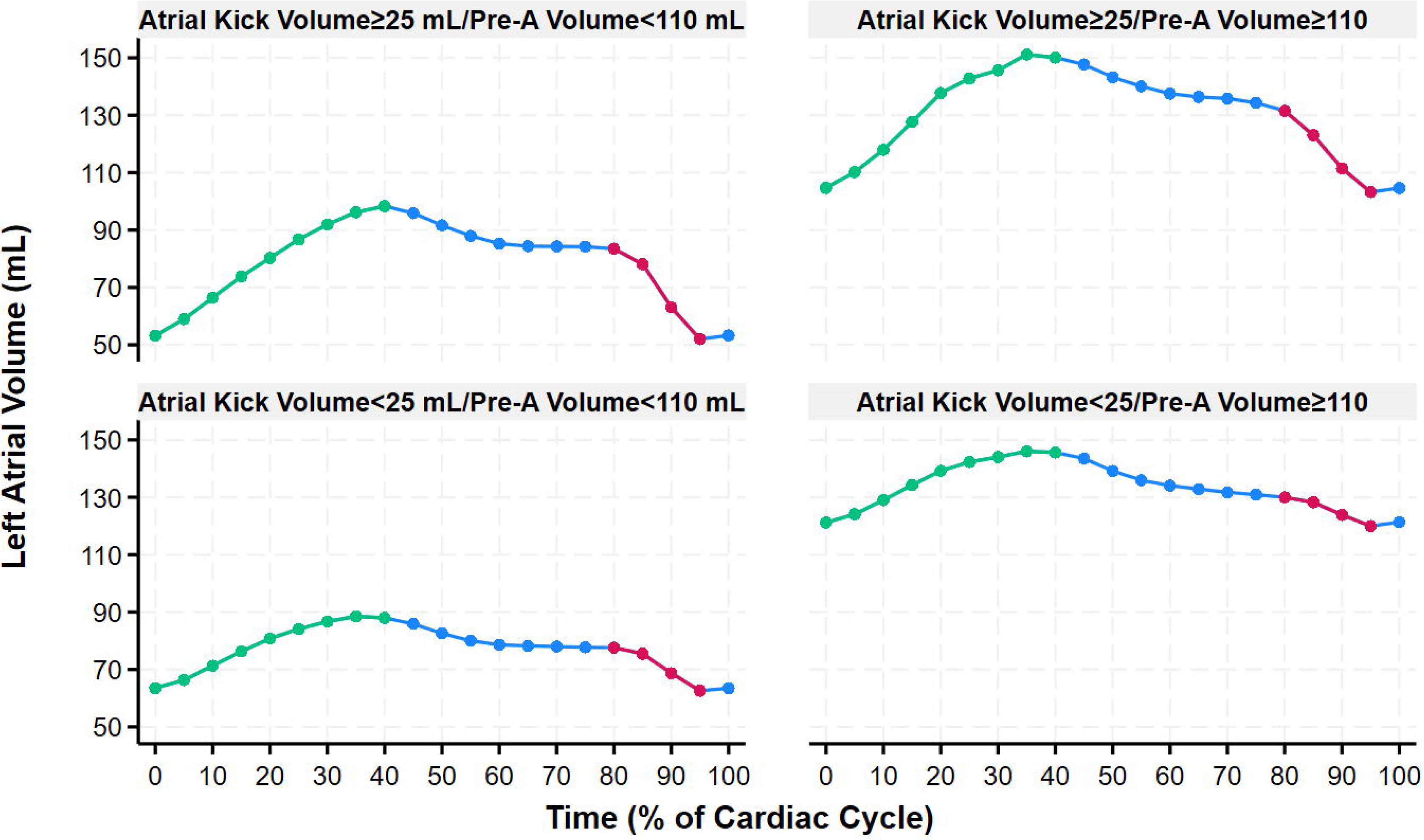
Left atrial volume-time curves according to pre-A and atrial kick volumes. The atrial booster is marked red. The left atrial filling period is marked green.

#### Relationship between LA booster function and reservoir function

There was a strong positive association between reservoir volume and LA booster volume (r=0.77, *P*<0.001; Figure 3), indicating that ∼60% of the variability in LA reservoir volume is explained by LA booster volume. In the subset of patients with flow data (n=74), there was a significant inverse correlation between the slope of the LA booster flow rate and the slope of LA reservoir filling rate during ventricular systole (Pearson correlation-0.79, *P*<0.001; Figure 4).

**Figure 3:**
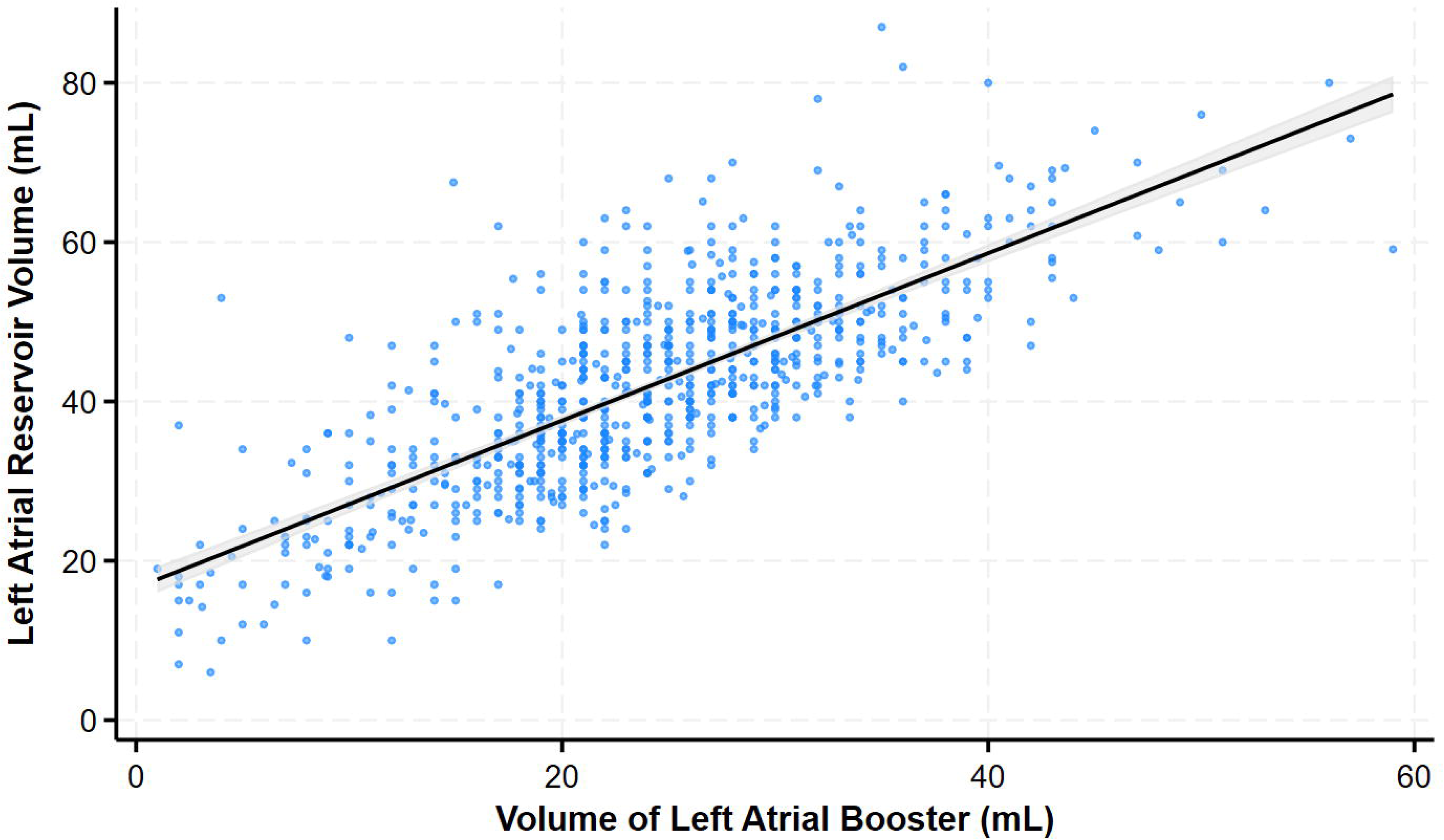
Relationship between the LA booster volume and the reservoir volume.

**Figure 4:**
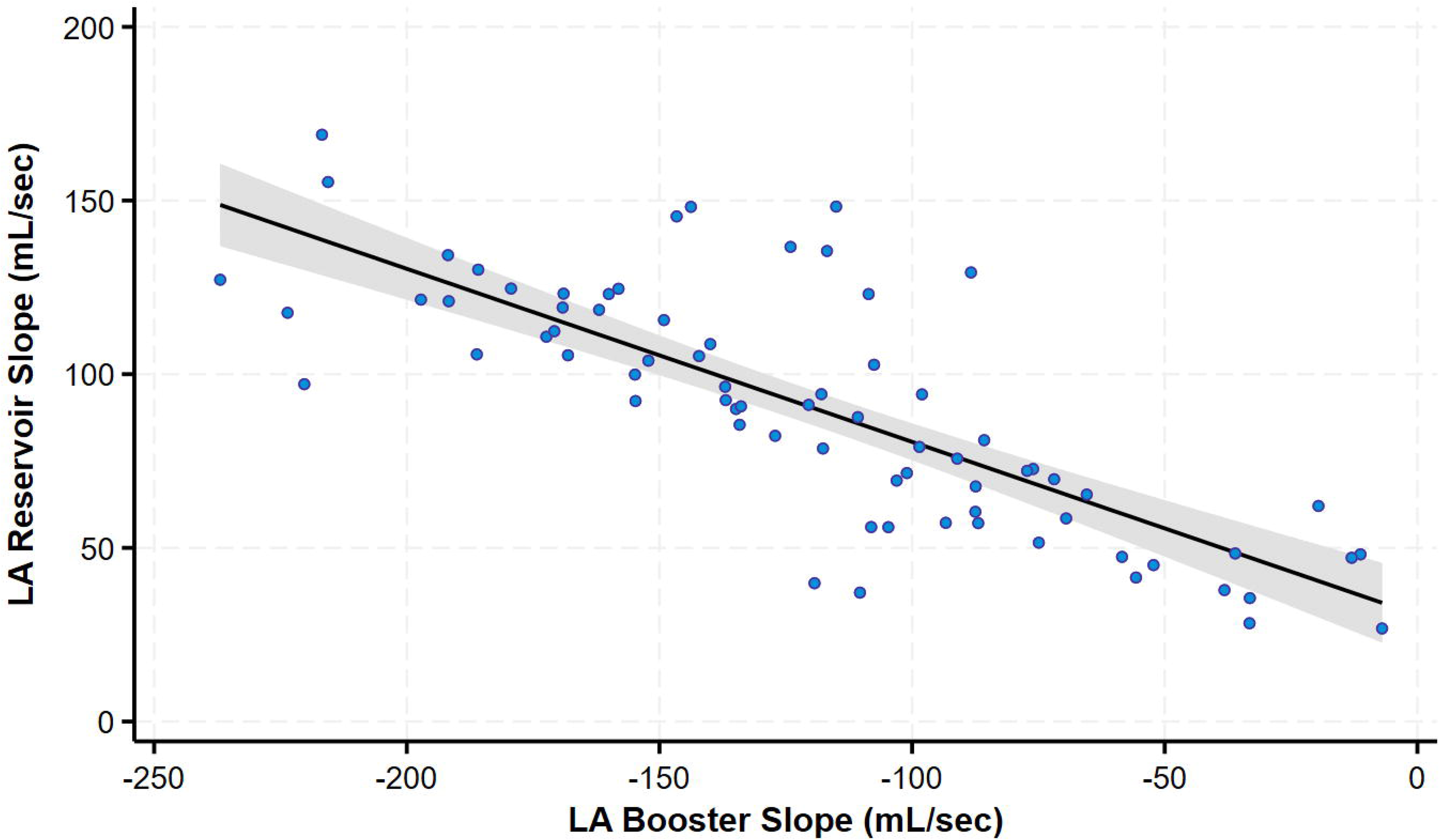
Relationship between the estimated LA booster flow rate and estimated LA filling rate in systole (n=74).

### Relationship between LA enlargement and LA pre-A volume

We next analyzed the determinants of LA pre-A volume. In contrast to what may be expected, a decrease in early passive emptying volume was not related to an increased pre-A volume. The LV early filling volume remained within a narrow range in the whole study population both as an absolute volume (mean 17.0 mL, 95% CI 16.5 to 17.5 mL) and as a fraction of LA volume (mean 16.1%, 95% CI 15.6 to 16.6%). Consequently, as can be seen in Figure 1A, LA passive emptying volume had no correlation with LA pre-A (Figure 5A). By contrast, LA maximal volume at end-systole strongly correlated with LA pre-A volume (Figure 5B). Linear regression demonstrated that for every 1 mL increase in maximum LA volume, the LA pre-A volume increased by 0.92 mL (95% CI 0.90 to 0.94 mL; *P*<0.0001), demonstrating that most of the increase in LA volume translated into an increase in LA Pre-A volume irrespective of the volume transferred to the LV in early diastole.

**Figure 5:**
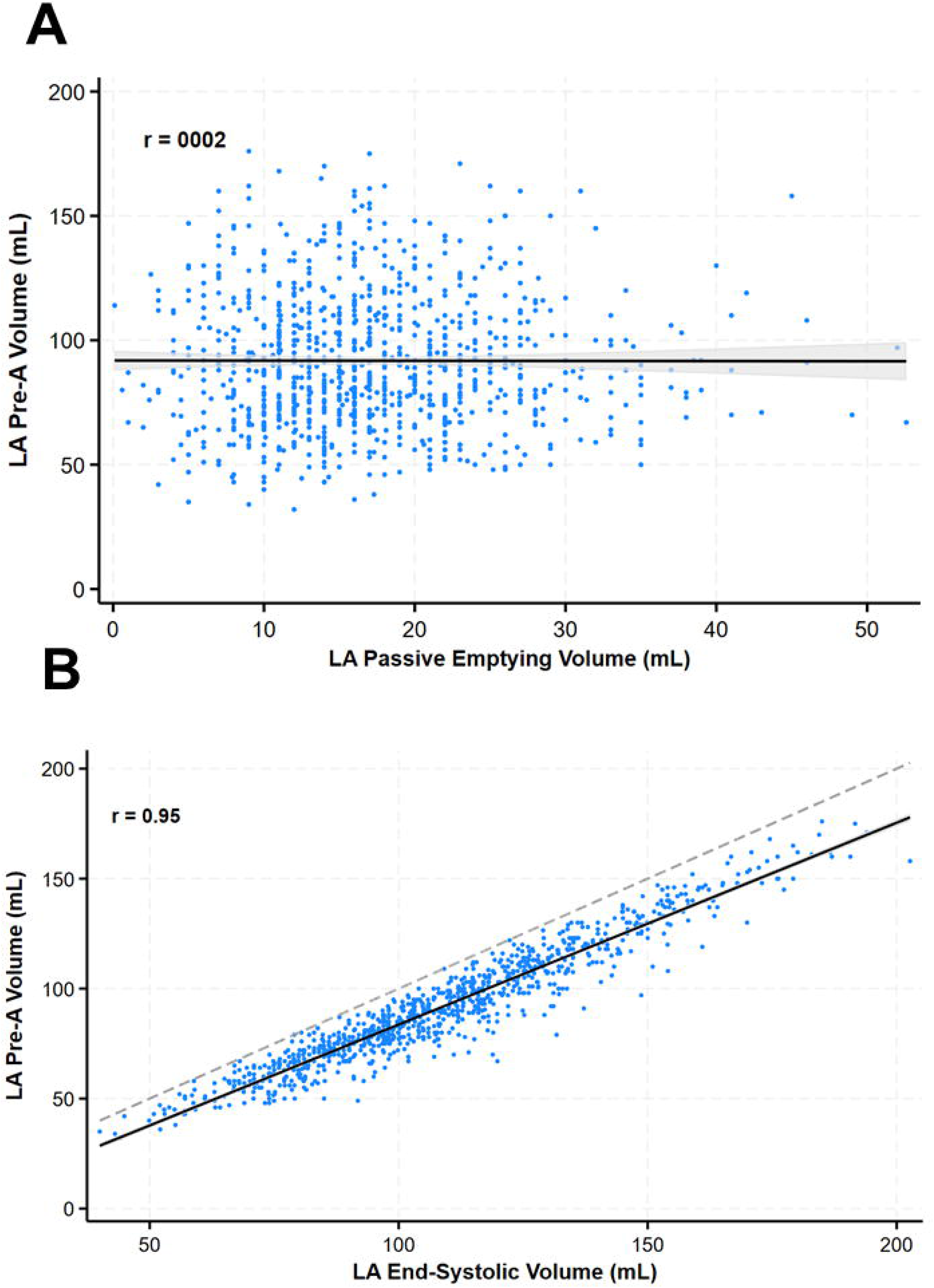
A. Relationship between LV filling in early diastole (LA early passive emptying volume) and LA pre-A volume. **B.** Relationship between maximal LA volume at end-systole and LA pre-A volume. The dashed line represents the line of identity between these measurements. The regression line approximates the line of identity (slope=0.92) with a proportional downward shift.

#### Clinical outcomes

Over a median follow-up of 3.9 years (IQR 3.8–4.1), the primary composite endpoint occurred in 175 patients (17.9 %). The Kaplan–Meier estimates of incident heart failure, stroke or cardiovascular mortality at 4 years (Figure 6) were lowest in group 2 patients with normal LAV**_pre-A_** and atrial kick volume (12.0% [95% CI 8.8–16.2%]) followed by group 1 patients with normal LAV**_pre-A_** and low atrial kick volume (17.4% [95% CI 13.3–21.8%]). Event rates increased to 30.1% (95% CI 22.2–41.3%) in group 3 patients with increased LAV**_pre-A_**and normal atrial kick volume and to 43.2% (95% CI 33.6–54.2%) in group 4 patients with increased LAV**_pre-A_**and low atrial kick volume (Figure 6).

**Figure 6:**
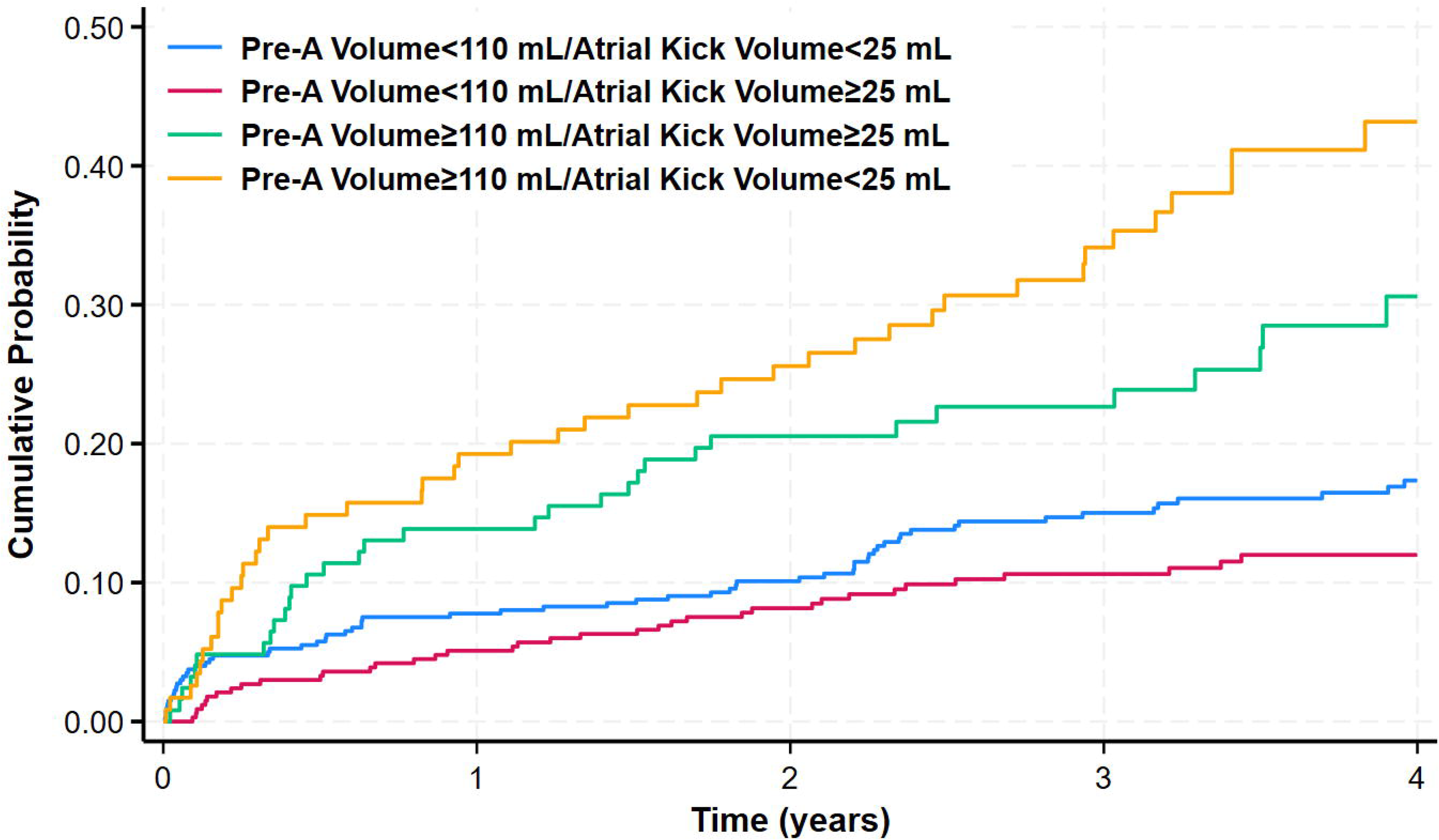
Kaplan–Meier estimates of incident heart failure or cardiovascular mortality according to LA volume pre-A wave and volume of the atrial kick.

Table 2 shows the results of Cox proportional hazards models for the composite endpoint. Compared with Group 2 with normal LAV**_pre-A_** and atrial kick volume as the reference group, there was a trend for increased risk in Group 1 patients with normal LAV**_pre-A_** and low atrial kick volume. Group 3 patients with increased LAV**_pre-A_** and normal atrial kick volume were also at increased risk with the highest risk in in Group 4 patients with increased LAV**_pre-A_** and low atrial kick volume.

## Discussion

In the present study, we characterized LA booster function in relation to LA remodeling using CTCA-derived volumetric measurements. With LA remodeling, the increase in LA volume predominantly translates into an increased LA pre-A volume, which may serve to preserve the atrial contribution to LV filling. However, the functional response to an increase in LA pre-A volume (atrial booster preload) varied substantially among patients. In some, LA remodeling was associated with a robust augmentation of booster function, reflected by maintained or increased LA active volume transfer to the LV. In others, LA booster function failed to rise despite substantial increases in LA pre-A volume, indicating impaired LA contractile reserve. LA systolic dysfunction strongly influenced overall LA performance through a concomitant reduction in reservoir function. Importantly, distinct patterns of functional remodeling were associated with divergent cardiovascular outcomes.

### LA enlargement as a compensatory mechanism

LA enlargement commonly accompanies cardiac diseases and generally reflects chronic elevation of left ventricular (LV) filling pressures (22). The ability of the LA to compensate for increased preload is critical for normal LA function. However, the impact of LA remodeling (i.e., increased LA volume) on overall LA function remains debated, particularly regarding whether it represents a compensatory or maladaptive response (3,17,23). Although LA enlargement can augment reservoir volume, such adaptive remodeling is primarily observed with physiological stimuli such as exercise training (24). In contrast, in pathological states, increased LAVI is often associated with impaired reservoir function (19,25,26).

Similar to the LV, LA enlargement requires higher preload to sustain effective booster function. However, in contrast to the LV which contracts at its maximal volume, LA systole occurs at a sub-maximal volume and pressure as some of the volume is transferred to the LV prior to atrial contraction (early passive filling). The current paradigm based on echocardiographic studies of mitral inflow suggest that reduced passive emptying of the LA leads to higher LA volume and pressure at the onset of atrial systole, thereby augmenting active emptying (3,23). In the present study, LA enlargement was accompanied by only minor changes in the volume transferred from LA to LV during early diastole, indicating that the reduced early emptying that frequently occurs with LA enlargement contributes little to increase LA preload. Instead, the rise in total LA volume is translated directly into increased LA pre-A volume, thereby enhancing the booster contribution to LV filling. Thus, LA enlargement primarily serves as a compensatory mechanism to augment booster function provided that LA contractility is not impaired. Indeed, LA enlargement was observed in nearly all patients with enhanced LA booster performance.

### LA booster function modulates reservoir function

Loss of LA contractile function was closely associated with a reduction in reservoir function, as reflected by the correlation between LA booster performance and the rate of LA filling during systole. This relationship likely stems from the influence of LA contraction on subsequent atrial relaxation, which facilitates early reservoir filling from the pulmonary veins. To the best of our knowledge, this study is the first to demonstrate this relationship in humans. These findings aligns with experimental pig model demonstrating that early LA reservoir filling depends on relaxation following the preceding contraction (27). However, impaired LA compliance and reservoir function may also develop in parallel with reduced booster function, driven by shared pathological processes such as LA cardiomyocyte hypertrophy and interstitial fibrosis. Conversely, an adaptive increase in LA booster function in response to LA enlargement can also preserve reservoir function and thereby maintain overall LA performance.

### The LA pre-A (preload) atrial booster relationship

Experimental studies in isolated rabbit left atria (28,29), and in humans (30) have demonstrated that the Frank–Starling mechanism does operate within the LA, whereby increased preload augments LA booster function. However, limited data are available in large patient populations or on its relationship to overall LA function or dysfunction.

Herein, we examined the relationship between LA pre-A volume—a measure of preload—and LA systolic stroke volume, analogous to the atrial Frank–Starling relationship. The resulting curve allows straightforward identification of patients with overt LA dysfunction, characterized by impaired booster function despite increased preload. Conversely, the coexistence of increased LA pre-A volume with preserved LA systolic function likely represents a compensated state where increased preload successfully recruits contractile reserve. This compensation is partial in some patients since overall LA performance remains mildly compromised and may readily progress to overt LA failure. Thus, we propose that the “LA pre-A volume versus LA systolic stroke volume curve” can serve as a functional framework to define LA failure.

Of note, among patients with LA pre-A volume < 110 mL, patients in group 1 had lower Global LA function, lower reservoir function and worse booster function. This group probably contains a mixture of patients with normal LA function and patients with (14).

LA contraction is modulated by 4 principal factors: (1) precontraction volume, a surrogate of precontraction fiber length; (2) the inotropic state; (3) LV afterload and (4) the degree of LA dysfunction (2,23,24,31). Variability in these determinants likely explains the heterogeneous functional response to LA remodeling observed among patients.

Failure of the LA booster pump may arise from increased workload imposed on the LA myocardium by elevated LV diastolic wall stress, which over time can lead to intrinsic atrial dysfunction. Additionally, afterload-independent mechanisms may contribute to progressive LA impairment. LA fibrosis, often secondary to risk factors such as obesity and hypertension, represents a central feature of the LA dysfunction phenotype (32).

### LA dysfunction and clinical outcomes

LA dysfunction is associated with increased risk of heart failure (33) and embolic stroke (34,35). In our cohort, patients demonstrating an apparently appropriate response to increased LA pre-A volume nonetheless remained at high cardiovascular risk. This phase may represent a transient compensatory state that rapidly progresses to LA contractile failure. Moreover, since conduit flow can maintain LV filling even in advanced LA dysfunction (17), the association between LA booster function and clinical outcomes may not solely reflect intrinsic LA mechanics, but also concomitant LV diastolic dysfunction. The poorest outcomes, however, were observed in patients with overt LA failure, defined by markedly increased LA pre-A volume accompanied by impaired booster function.

## Conclusion

Increase in LA volume predominantly serves to increase LA Pre-A volume, serving to sustain LA booster function. LA failure can be defined as reduced booster function despite an elevated LA pre-A volume. Because LA systolic dysfunction directly impairs reservoir function, it contributes to global LA impairment. Distinct patterns of LA functional remodeling were associated with divergent cardiovascular outcomes, underscoring the clinical relevance of characterizing LA mechanics beyond size alone.

## Data Availability

All data produced in the present study are available upon reasonable request to the authors

