## Supplemental File for "Characterizing Left Atrial Failure via the Atrial Booster Preload-Performance Relationship"

**Supplemental Figure 1:** Nonparametric regression of the relationship between pre-A volume and atrial kick volume.

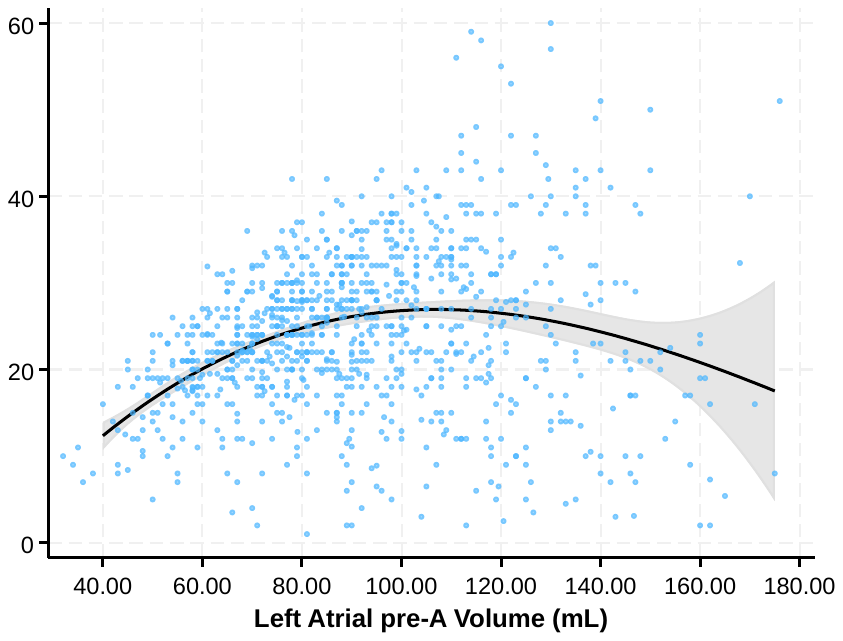

**Supplemental Figure 2:** Quantile regression of the relationship between pre-A volume and atrial kick volume. The black line represents the median fit. Gray lies represent the 5^th^ and 95th percentiles.

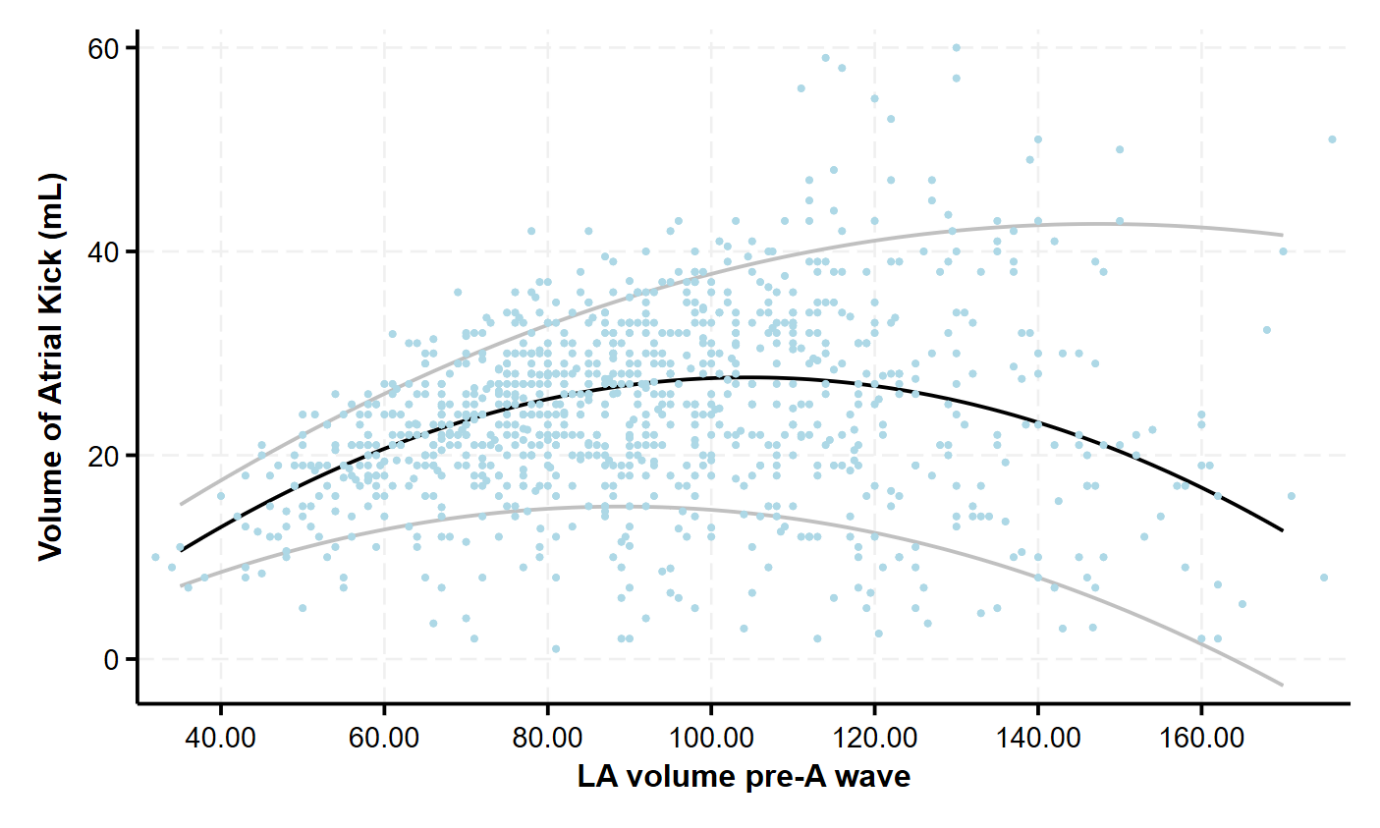

**Supplemental Figure 3:** Relationship between LA volume pre-A wave and the volume of the atrial kick in patients with preserved, and reduced or mid-range ejection fraction.

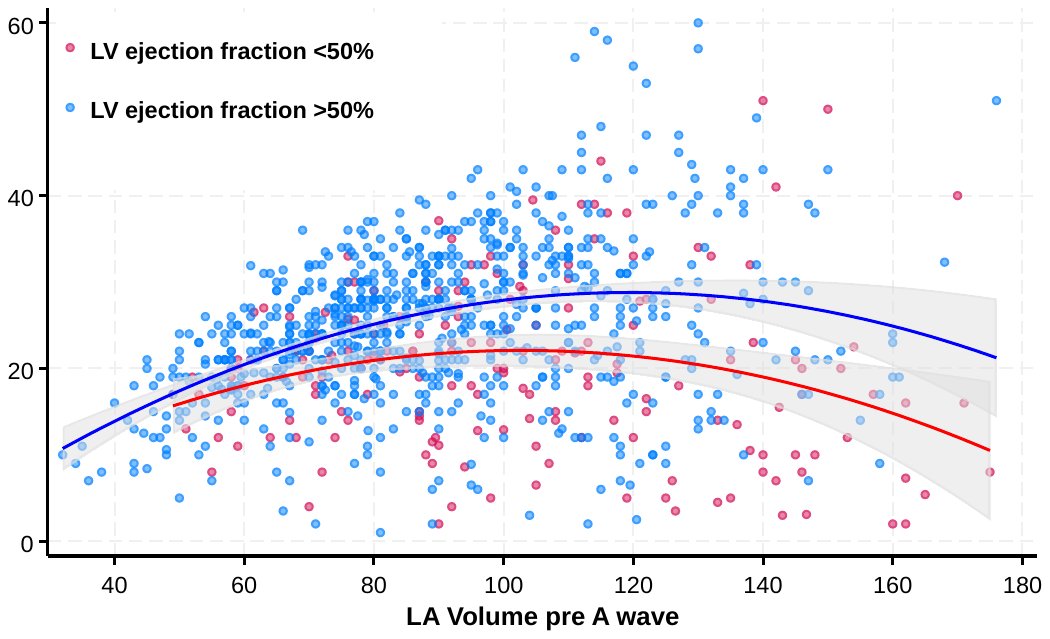

**Supplemental Figure 4:** Relationship between LA volume pre-A wave and the volume of the atrial kick in females and males.

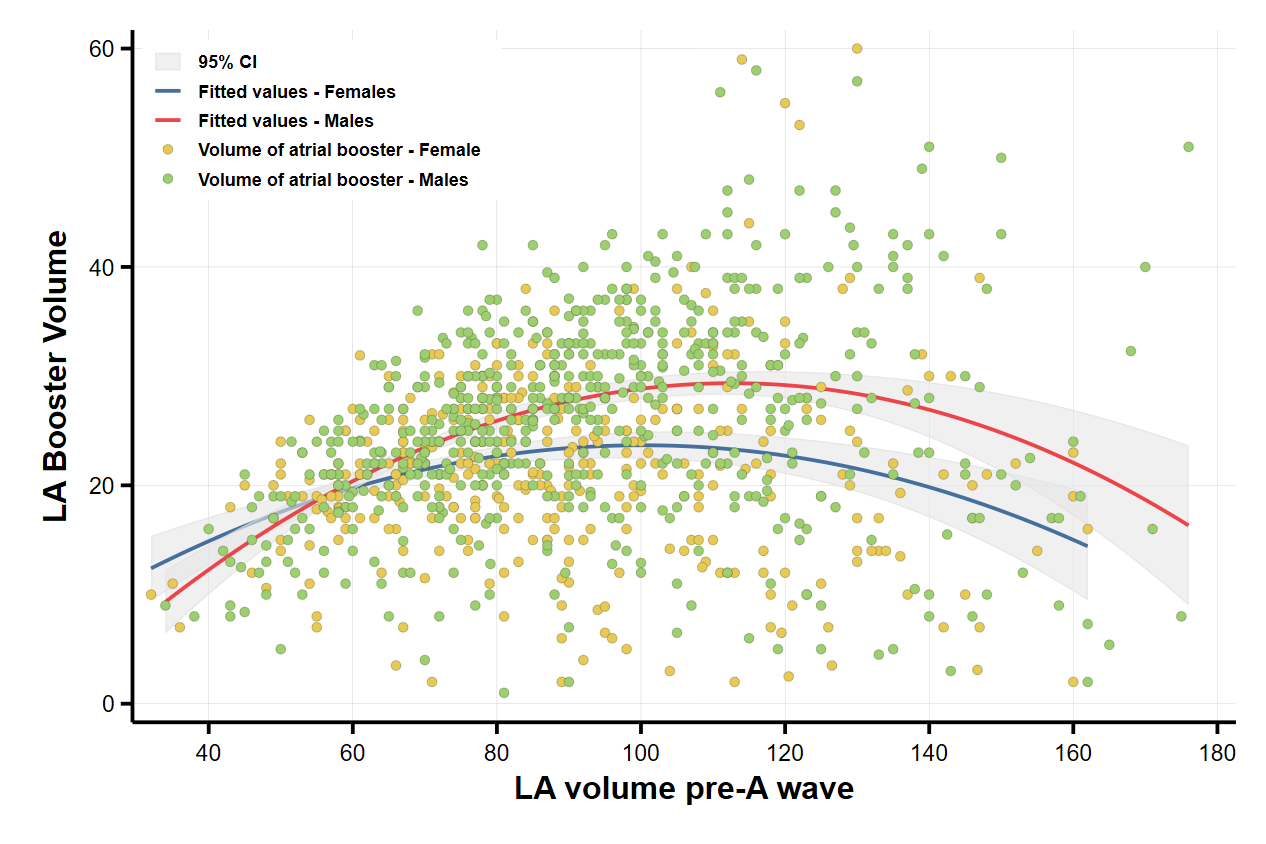
